# Kynurenic acid underlies sex-specific immune responses to COVID-19

**DOI:** 10.1101/2020.09.06.20189159

**Authors:** Yuping Cai, Daniel J. Kim, Takehiro Takahashi, David I. Broadhurst, Shuangge Ma, Nicholas J.W. Rattray, Arnau Casanovas-Massana, Benjamin Israelow, Jon Klein, Carolina Lucas, Tianyang Mao, Adam J. Moore, M. Catherine Muenker, Jieun Oh, Julio Silva, Patrick Wong, Yale IMPACT Research team, Albert I. Ko, Sajid A. Khan, Akiko Iwasaki, Caroline H. Johnson

## Abstract

Coronavirus disease-2019 (COVID-19) has poorer clinical outcomes in males compared to females, and immune responses underlie these sex-related differences in disease trajectory. As immune responses are in part regulated by metabolites, we examined whether the serum metabolome has sex-specificity for immune responses in COVID-19. In males with COVID-19, kynurenic acid (KA) and a high KA to kynurenine (K) ratio was positively correlated with age, inflammatory cytokines, and chemokines and was negatively correlated with T cell responses, revealing that KA production is linked to immune responses in males. Males that clinically deteriorated had a higher KA:K ratio than those that stabilized. In females with COVID-19, this ratio positively correlated with T cell responses and did not correlate with age or clinical severity. KA is known to inhibit glutamate release, and we observed that serum glutamate is lower in patients that deteriorate from COVID-19 compared to those that stabilize, and correlates with immune responses. Analysis of Genotype-Tissue Expression (GTEx) data revealed that expression of kynurenine aminotransferase, which regulates KA production, correlates most strongly with cytokine levels and aryl hydrocarbon receptor activation in older males. This study reveals that KA has a sex-specific link to immune responses and clinical outcomes, in COVID-19 infection.

## Main

Sex-related differences in coronavirus disease-2019 (COVID-19) severity and morbidity exist, with the male sex being a risk factor^1^; male COVID-19 patients have an increased risk of admission (OR 1.68, 95%CI = 1.45-1.90) and in-hospital mortality (OR 1.87, 95%CI = 1.33-2.63)^1^. It was recently shown that hospitalized patients with moderate SARS-CoV-2 infection have elevated levels of certain inflammatory cytokines and chemokines, and sex-differences exist in these immune responses^2^. Furthermore, across all ages, female patients at baseline had a more robust T cell activation than males. Loss of T cell activation was correlated with older age in males, and this poorer T cell response was correlated with worse disease outcomes in males only^2^. Therefore, males and females have clear differences in COVID-19 immune responses that correlate with clinical course.

Since immune responses are regulated, in part, by metabolites, it is possible that sex-related differences in metabolism could affect the host immune response to SARS-CoV-2 infection. For instance, specific metabolites are required for macrophage, neutrophil, and T cell functions, enhancing glycolytic and fatty acid synthesis pathways in these cells^3^. Conversely, immune stimulation can also elicit metabolic reprograming in cells, thereby affecting disease trajectory by altering metabolite abundance^4^. In addition to the metabolic requirements of the host immune system, viruses also require host-derived metabolites and lipids^5^. Thus, utilization of metabolic substrates for viral replication could affect metabolite availability required for immune responses.

### Metabolites correlate with COVID-19

To address how metabolites might mediate the sex-related differences in COVID-19 immune response, we first used an untargeted metabolomics workflow with multivariable logistic regression to identify serum metabolites associated with COVID-19. Serum samples were collected from COVID-19 patients (n = 22 females and n = 17 males) on the day of enrollment into the IMPACT study at Yale New Haven Hospital (CT, USA). Samples were taken from patients 3-7 days after hospital admission after confirmation of COVID-19 infection who (1) were not immediately triaged to the intensive care unit, (2) had not received tocilizumab, and (3) had not received high dose corticosteroids (Cohort A described in Takahashi et al.^2^). Uninfected healthcare worker (HCW) controls (n = 10 females and n = 10 males) were included in the analysis. There was a statistically significant difference in age between the COVID-19 patients and HCWs, which was adjusted for in our models (**Extended Data Table 1**). We first carried out metabolite identification on detected signals that were present in the serum metabolome of quality control samples pooled from both COVID-19 patients and HCWs. We positively identified 75 metabolites with the highest confidence (**Extended Data Table 2**). Regression analysis revealed that 17 metabolites were associated with COVID-19 status after adjustment for age, BMI, sex, and multiple comparisons (**Extended Data Table 3**). Glutamate, cysteine-S-sulfate, palmitoleic acid, arachidonic acid, lysophosphatidylethanolamine (LPE) (22:6), uracil and myristic acid were positively associated with COVID-19. Whereas glutamine, 3-methylxanthine, tryptophan, proline, citrulline, homoserine, 2,3-dihydroxybenzoic acid, lysophosphatidic acid (LPA) (18:2), LPA (20:2), and lysophosphatidylcholine (14:0) were negatively associated with COVID-19.

### Metabolites correlate with immune response in a sex-specific manner

Next, we examined how the 75 positively identified serum metabolites from both COVID-19 patients and HCWs might correlate with immune markers (cytokines and chemokines levels in plasma, T cells, B cells, NK-T cells, NK cells, monocytes, macrophages, and dendritic cells in peripheral blood mononuclear cells (PMBCs)) that were previously measured from the same individuals^2^ (**Fig. 1**). In COVID-19 patients, we observed 36 correlations between immune markers and metabolites with |R|>0.5 and *p* value< 0.05 (**Extended Data Fig. 1,Supplementary Table 1**). However, after stratification by sex, additional correlations were observed between metabolites and immune markers for each sex independently, suggesting that sex-specific immune responses could be linked to metabolism (**Extended Data Fig. 1, Supplementary Table 2**).

**Fig. 1.**
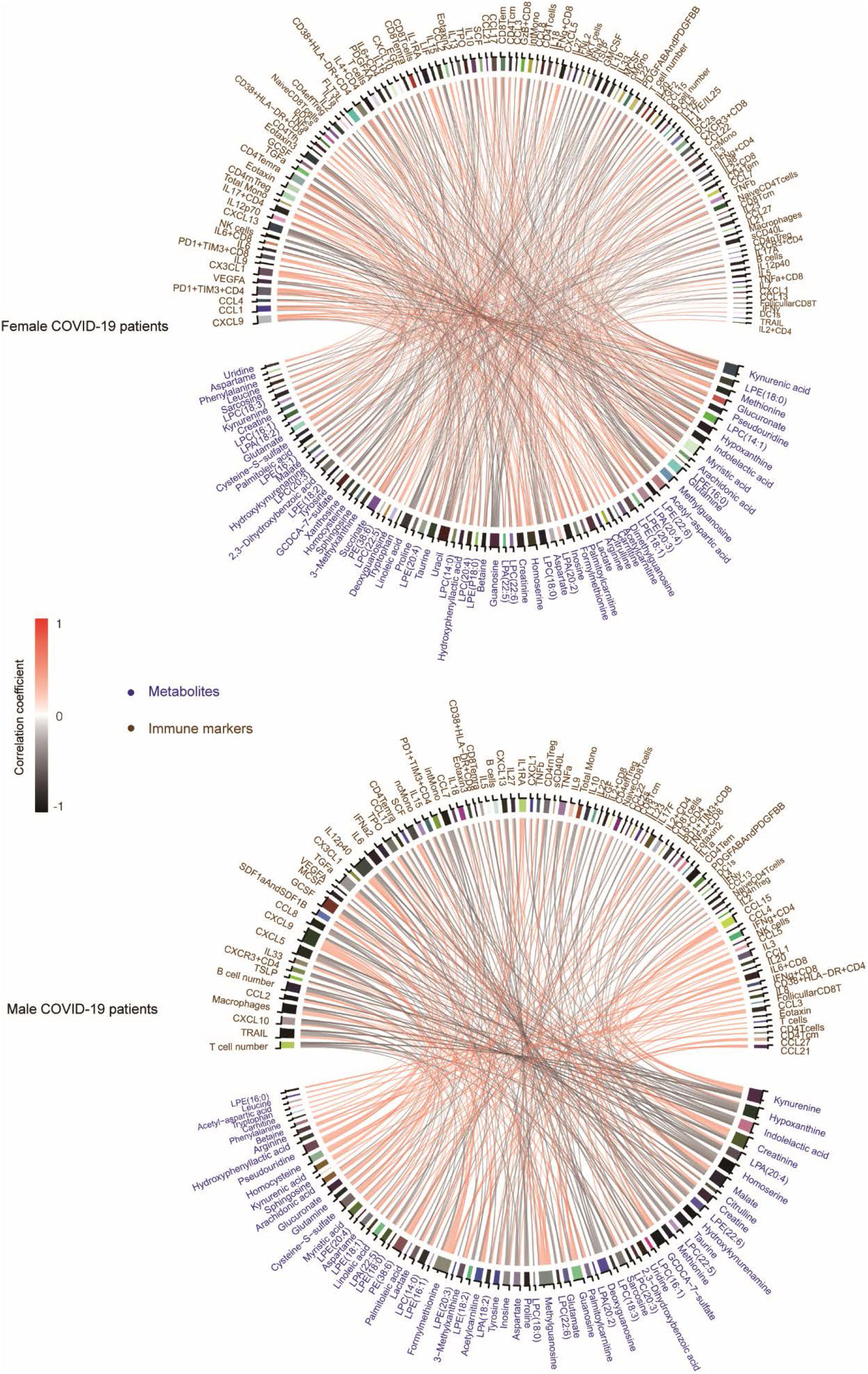
Chord diagram of correlations between metabolites and immune markers in COVID-19 patients. Spearman correlations > 0.5 or < −0.5 are displayed and with p< 0.05.

Further examination revealed that kynurenic acid (KA), an endogenous ligand of the aryl hydrocarbon receptor (AhR) that regulates immunes responses^6^, had positive correlations with immune markers (**Fig. 1**). Many of these positive correlations were observed in male patients including IL1RA, IL6, IL10, TNFα, M-CSF, SCF, CX3CL1, CXCL9, CXCL13, CCL1, CCL21, and CCL22. In addition, KA in males was negatively associated with T cell number, naïve CD8 T cells, CD4 effector memory (CD4Tem), and CD8 effector memory (CD8Tem) T cells (**Figs. 2a, 2b**). In female patients, KA was positively associated only with IL12p40, CCL3, CXCL9, and SCF (**Fig. 2b**). In summary, sex-specific differences in correlations between metabolites and immune responses were observed in COVID-19 patients, wherein KA had the most prominent connection to immune response in males.

**Fig. 2.**
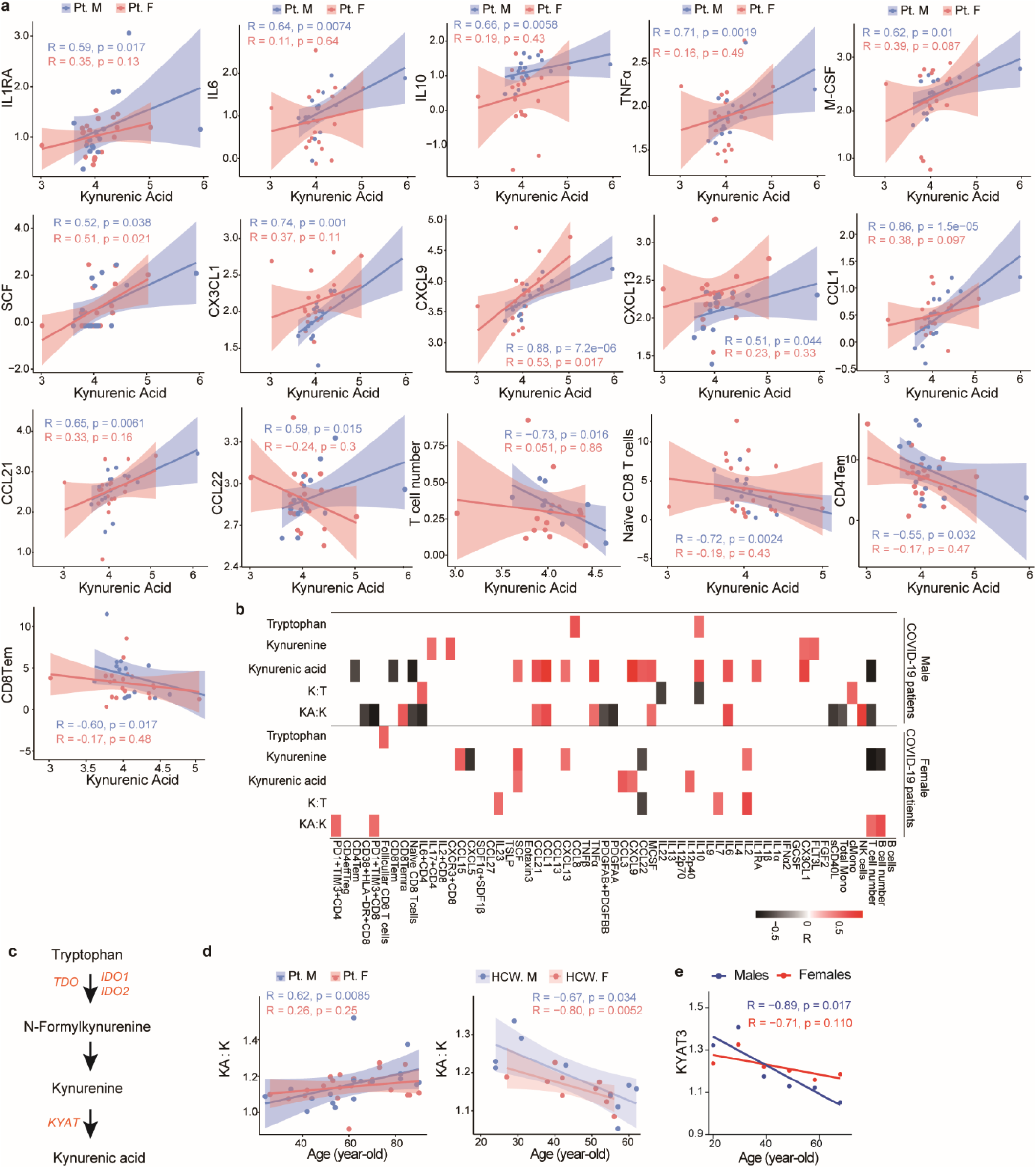
Tryptophan pathway metabolites and immune responses. **a,** Correlation between kynurenic acid (KA) and immune markers in males with COVID-19 (Pt. M, n = 17) and females with COVID-19 (Pt. F, n = 22). 95% confidence intervals (CIs) for the correlation coefficients are indicated as shaded areas colored according to patient sex. **b**, Tryptophan (T) metabolism pathway schematic. **c**, Heatmap showing correlation between tryptophan metabolites and immune markers in males and females with COVID-19. Spearman correlations > 0.5 or < –0.5 are displayed, p< 0.05. **d**, Correlation between age and KA:kynurenine (K) ratio in patients with COVID-19 and healthcare workers (HCWs). **e**, Correlation between *KYAT3* (expression averaged within each age group) and age in Genotype-Tissue Expression (GTEx) samples (n = 729 males, 1914 females). Metabolites are displayed as ion intensity log10 transformed, cytokines and chemokines are pg/mL log10 transformed, T cell subsets are % in CD3 T cells, T cell number are 10^6 cells/mL, PBMCs are % in live PBMCs.

**Fig. 3.**
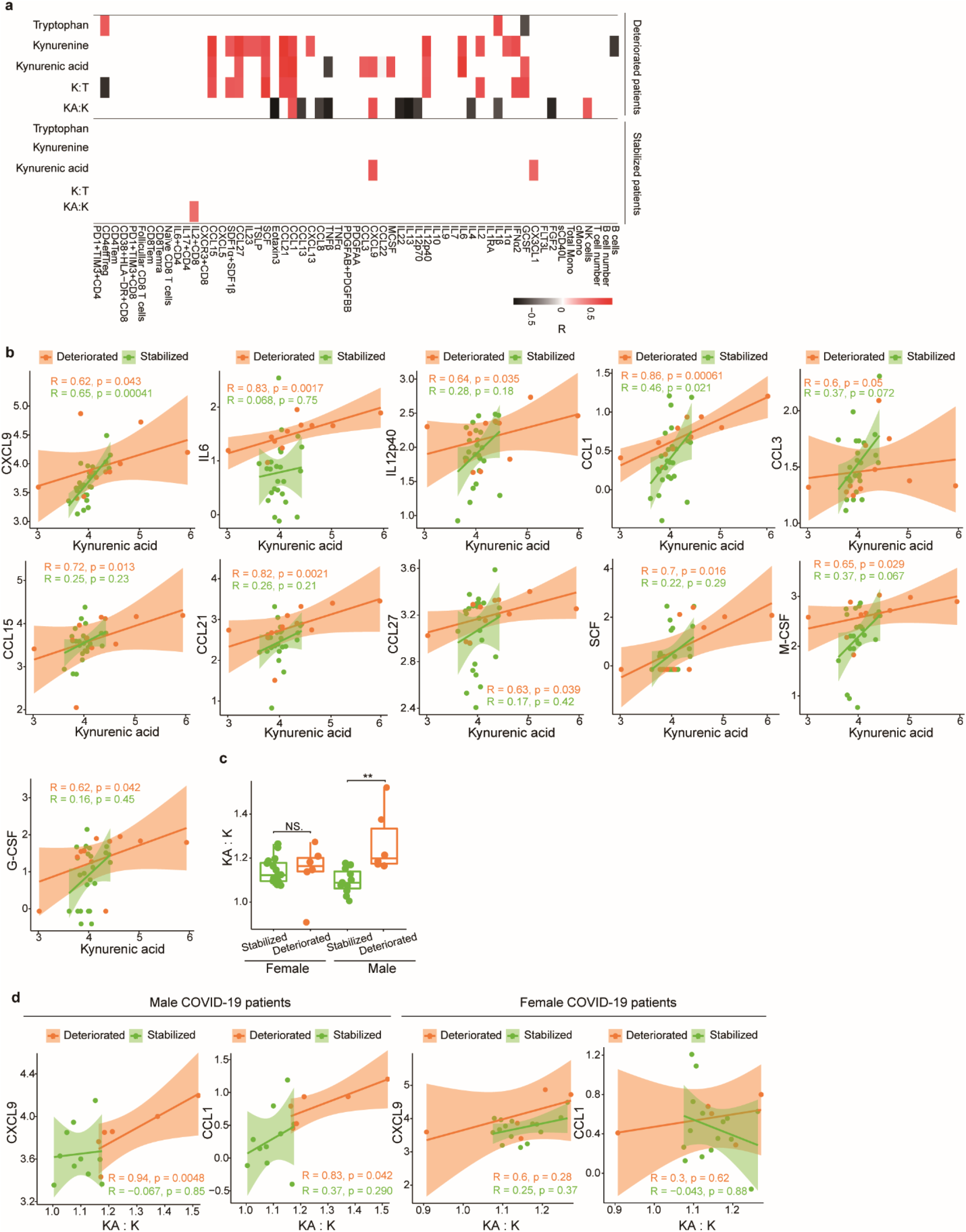
Tryptophan metabolites, immune markers and disease severity. **a**, Heatmap of correlations between metabolites in the tryptophan pathway and immune markers by disease severity. Spearman correlations > 0.5 or < –0.5 are displayed with a p< 0.05. **b**, Correlation between kynurenic acid (KA) and immune markers by disease severity. 95% confidence intervals (CIs) for the correlation coefficients are indicated in shaded area colored according to disease progression status. **c**, Comparison of the ratio of KA:kynurenine (K) level by disease severity stratified by sex. Stabilized (females n = 16, males = 11), deteriorated (females n = 6, males n = 6). Nonparametric Kruskal–Wallis rank sum test with pairwise Wilcoxon Mann-Whitney U test, *p* values adjusted for false discovery rate (Benjamini-Hochberg). \*\**p*< 0.01, NS. not significant. **d**, Correlation between the ratio of KA:kynurenine (K) and CXCL9 and CCL1 stratified by disease severity and sex. Metabolites are displayed as ion intensity log10 transformed, cytokines and chemokines are pg/mL log10 transformed, T cell subsets are % in CD3 T cells, T cell number are 10^6 cells/mL, PBMCs are % in live PBMCs.

### Kynurenic acid is associated with a sex-specific immune response

To further understand the sex-specific correlation of KA to immune markers, we examined the relationship between KA and kynurenine (**Fig. 2b**). Kynurenine (K) is a product of tryptophan metabolism that is converted to KA by kynurenine aminotransferases (KATs), which are encoded by *KYAT* genes (**Fig. 2c**). Of note, tryptophan was inversely associated with COVID-19 disease, as noted in **Extended Data Table 3**. We examined the ratio of KA:K in patients with COVID-19 as a surrogate for KAT-mediated production of KA from K^7^. In males, we observed that a high KA:K was positively correlated with IL6, CCL1, CCL21, TNFα, M-CSF, NK cells, and CD8 terminally differentiated effector memory (Temra) T cells (**Fig. 2b**). A high KA:K was negatively correlated with sCD40L, PDGFAA, PDGFAB/BB, monocytes, PD1+TIM3+CD8 T cells, CD38+HLA-DR+CD8+ T cells, naïve T cells, and IL6^+^CD4 T cells (**Fig. 2c**). Of note, a high KA:K was positively correlated with T cell activation in females, but overall T cell numbers showed a negative correlation with the ratio of KA:K in males with COVID-19 (**Fig. 2c)**.

We also observed that KA:K and KA positively correlated with age in males with COVID-19 (**Fig. 2d, Extended Data Fig. 2a**). KA had a low positive correlation to age in females with COVID-19, but the ratio of KA:K was not correlated (**Extended Data Fig. 2a, Fig. 2d**). In HCWs, KA negatively correlated with age only in males (**Extended Data Fig. 2a**), while KA:K negatively correlated with age in both males and females (**Fig. 2d**).

Closer examination of other metabolites involved in K and KA metabolism revealed additional correlates of the cellular immune response during COVID-19. The microbial catabolite of tryptophan, indole-3-lactic acid^8^, was positively associated with IL4+CD4 and CD38+HLADR+CD8 cells in males (**Extended Data Fig. 2b**). In females, indole-3-lactic acid was negatively associated with plasma levels of G-CSF, M-CSF, and CXCL10; K was positively associated with IL2, CCL15, CXCL13, and SCF; and tryptophan was positively correlated with follicular CD8 T cells (**Fig. 2b**).

To evaluate whether the sex-specific association between KA and the immune response, which was observed in males with COVID-19, is a phenomenon also present in healthy individuals, we analyzed tissue-specific expression data from the Genotype-Tissue Expression (GTEx) Project^9^. While *KYAT* genes generally tended to have more positive correlations with cytokines in males compared to females, *KYAT3* had particularly stronger correlations in a subset of tissues (including the brain and colon), many of which are classically involved in COVID-19 (**Extended Data Fig. 3**). Within the brain, we found that these positive correlations with cytokines were specific to older males (aged > 60 years old) (**Extended Data Fig. 4a**). Because KA is a ligand for AhR which regulates immune responses and inflammation^6^, we analyzed AhR activation using a previously defined score^10^ and found that AhR activation correlates most positively with *KYAT3* expression in older males in brain and muscle, while closely correlating in colon (**Extended Data Fig. 4b**). Correlations in the brain became even more pronounced when analyzing only the AhR target gene *CYP1B1*, which is classically used to follow AhR activation in the brain (**Extended Data Fig. 4c**)^11^. We also show that *KYAT3* expression decreases with age in both males and females, which is consistent with the decreasing ratios of KA:K observed in HCW control samples (**Fig. 2e**). In summary, these data suggest that older males are uniquely sensitive to increases in KA levels, reacting disproportionately with increased levels of inflammatory cytokines, likely as part of a broader AhR activation.

**Fig. 4.**
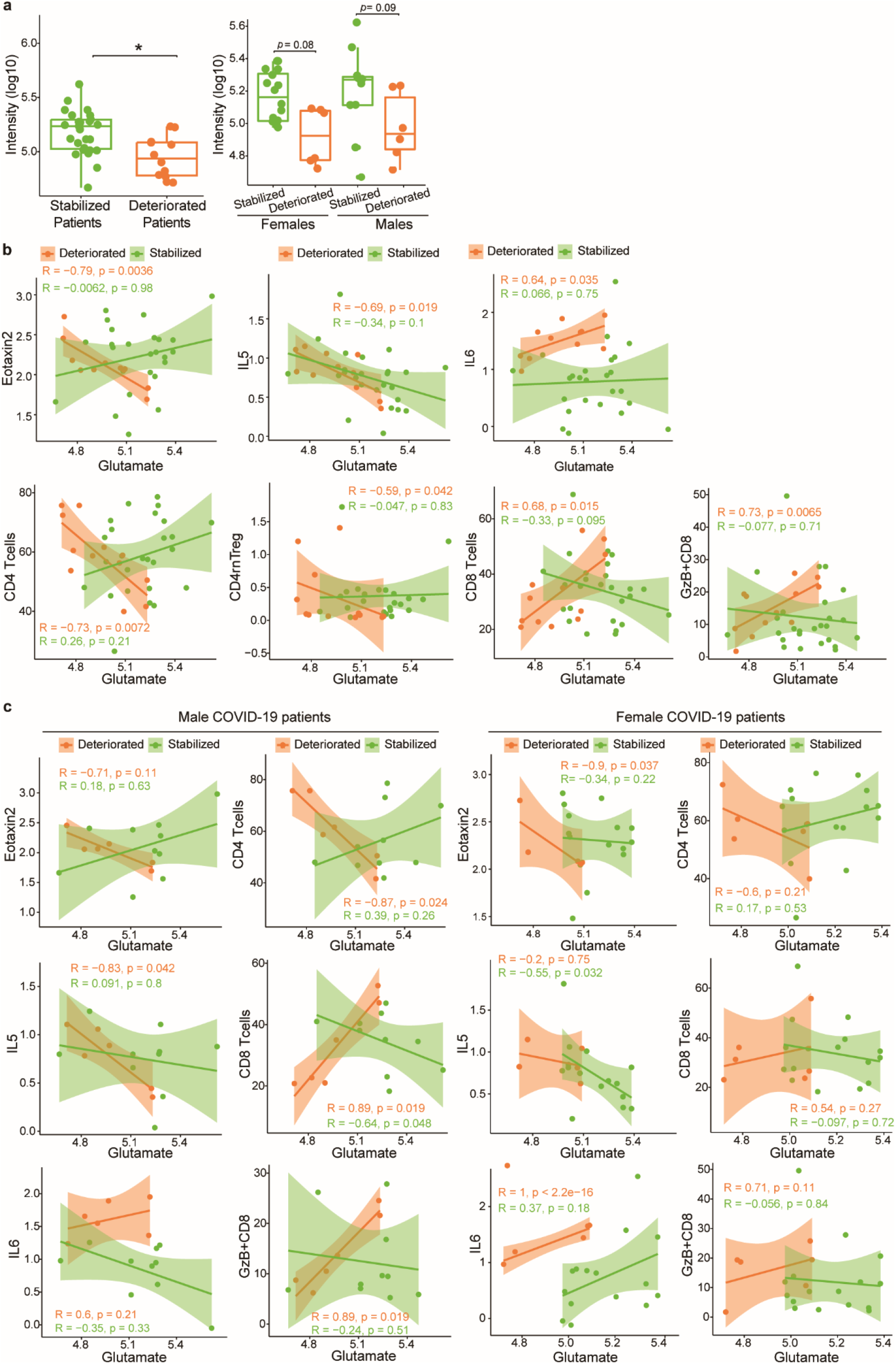
Glutamate, immune markers and disease severity. **a**, Comparison of glutamate levels in stablized patients and deteriorated patients (left panel) and stratified by sex (right panel). Stabilized patients (n = 27), deteriorated patients (n = 12), stabilized females (n = 16), deteriorated females (n = 6), stabilized males (n = 11), and deteriorated males (n = 6). Nonparametric Kruskal–Wallis rank sum test with pairwise Wilcoxon Mann-Whitney U test, *p* values adjusted for false discovery rates (FDR) (Benjamini-Hochberg). \*\**p*< 0.01, NS. not significant. **b**, Correlation between glutamate and Eotaxin2, IL5, IL6, CD4 T cells, CD4rnTreg cells, CD8 T cells and GzB+CD8 cells in stabilized patients and deteriorated patients. **c**, Correlation between glutamate and immune markers eotaxin2, IL5, IL6, CD4 T cells, CD8 T cells, GzB+CD8 cells, and IL6 in stabilized patients and deteriorated patients stratified by sex. 95% confidence intervals (CIs) for the correlation coefficients were indicated as the shadowed area colored according to progression status. Metabolites are displayed as ion intensity log10 transformed, cytokines and chemokines are pg/mL log10 transformed, T cell subsets are % in CD3 T cells, T cell number are 10^6 cells/mL, PBMCs are % in live PBMCs.

### Kynurenic acid correlates with disease severity in a sex-specific manner

As KA levels correlate with numerous immune markers, and these immune markers have been previously linked disease progression^2^, we examined whether KA was directly associated with disease severity. We used previously defined clinical scores to classify disease severity in COVID-19 patients as deteriorated (males n = 6, females n = 6) or stabilized (males n = 11, females n = 16)^2^. KA levels were not significantly different between deteriorated and stabilized patients, or after additional stratification by sex. However, KA was positively correlated with CXCL9, IL6, IL12p40, CCL1, CCL3, CCL15, CCL21, CCL27, SCF, M-CSF, and G-CSF in patients that deteriorated with COVID-19 (|R|>0.5 and *p* value< 0.05). In stabilized patients, KA was also positively correlated with CXCL9 and CX3CL1 (**Figs. 3a, b**). We further examined whether KA:K was correlated with disease severity by sex. Males who deteriorated had a significantly higher KA:K compared to those that stabilized, whereas there was no difference in KA:K between females based on clinical course (**Fig. 3c**). Furthermore, a high KA:K was positively correlated with CXCL9 and CCL1 in males that deteriorated, but this correlation was not seen in patients that stabilized, or in females (**Fig. 3d**).

We also examined whether any of the 17 metabolites associated with COVID-19 status (**Extended Data Table 3**) were correlated with disease severity. We observed that only glutamate was associated with disease trajectory, and a significantly higher glutamate level was observed in stabilized patients compared to those that deteriorated (**Fig. 4a)**; incidentally, KA is a glutamate receptor antagonist, thus high KA production could inhibit glutamate release^12^. Correlation analysis revealed that Eotaxin2, IL5, CD4 T cells, CD4rnTreg negatively correlated with glutamate in deteriorated patients, whereas IL6, CD8 T cells and GzB+CD8 T cells positively correlate (**Fig. 4b)**. Further stratification by sex, showed a similar trend where stabilized patients had higher levels of glutamate than deteriorated patients within each sex group (**Fig. 4a)**. Correlation analysis of immune markers with glutamate by sex revealed a negative correlation to Eotaxin2 and positive correlation to IL6, only in females that deteriorate. CD4 T cells negatively correlate and GzB+CD8 T cells positively correlate with glutamate, only in males that deteriorate. IL5 negatively correlates with glutamate in males who deteriorate whereas it negatively correlates with glutamate in females that stabilize. CD8 T cells positively correlate with glutamate in males that deteriorate and negatively correlate in males that stabilize (**Fig. 4c**). These data suggest that low levels of glutamate may contribute to poorer disease progression in patients with COVID-19 by regulating immune responses. In addition, a high KA:K is correlated to poorer prognosis only in male COVID-19 patients.

### Discussion

Patients with severe COVID-19 disease experience a “cytokine storm”, characterized by the elevation of pro-inflammatory cytokines and aggressive inflammatory response^13^, and sex-specificity in immune response has been previously reported that could underlie differences in clinical outcomes^2^. Our analysis of serum metabolites from COVID-19 patients, reveals that KA and the ratio of KA:K has a strong relationship to sex-specific immune response and clinical disease course. Importantly, a previous study revealed that serum metabolites in tryptophan and K metabolism correlate with IL6 in a sex-aggregated cohort of COVID-19 patients, but sex-specificity was not examined^14^. Our study shows that, in males, a high ratio of KA:K was positively correlated with increased levels of cytokines/chemokines and portends clinical deterioration. On the other hand, a negative association was observed with Eotaxin, sCD40L, PDGFs, and T cells, indicating that males with a high KA:K may have a poorer response to inflammation associated with COVID-19, including decreased eosinophil recruitment and T cell activation^15,16^. A previous study showed that PDGFs associate with better prognosis from COVID-19 if the patients have lower cytokine levels, supporting the association between a higher KA:K and poorer outcomes in males^17^. In females with COVID-19, a high KA:K positively correlated with a small number of cytokines and also T cell activation, but in contradistinction to males, high KA:K was not associated with disease severity. These results therefore support the role of K metabolism in sex-related differences previously reported in immune responses to COVID-19^2^.

Analyzing gene expression data from GTEx, we found that older males (but not females or younger males) appear to have exquisite sensitivity to changes in *KYAT* gene expression (which we used as a proxy for KA levels), whereby natural increases in *KYAT* expression are met with concomitant natural increases in tissue cytokine expression. It is worthwhile to note that the tissues exhibiting these sex-specific correlations – including brain, muscle, kidney, and colon – are those that are commonly implicated in symptoms of COVID-19 patients such as anosmia, myalgia, acute kidney injury, and gastrointestinal distress.

Given its role in regulating the immune system and inducing pro-inflammatory cytokines like IL6^6^, modulations in the AhR signaling pathway likely account for this differential response among older males. In support of this hypothesis, we show that AhR activation is most strongly associated with *KYAT3* expression in healthy older males. Also, studies have already shown that male rodents have a more toxic response to stereotypical AhR agonists like TCDD^18^. Furthermore, testosterone-mediated signaling is known to inhibit AhR activity^19^, and given the decreasing serum levels of testosterone seen in older males^20^, it seems plausible that healthy older males could be naturally susceptible to greater AhR activation by endogenous ligands.

In the context of COVID-19 infection, patients presented with elevated KAT activity (as suggested by high KA:K ratios), especially among deteriorating male patients. A recent study has demonstrated that a similar AhR induction occurs in the context of murine coronavirus infection, inducing IDO-2 expression^17^. Notably, two major risk factors for COVID-19, type 2 diabetes and obesity, have already been shown to have increased AhR ligand activity^21,22^. Such an influx of endogenous AhR ligands, combined with an already elevated susceptibility for AhR activation, therefore, would pose a significantly elevated risk of developing a cytokine storm, specifically in older male patients.

The analysis in this study also revealed discrete serum metabolites associated with COVID-19 that may account for some of the varying clinical outcomes in these patients. For instance, metabolites that were positively associated with COVID-19 (**Extended Data Table 3**) have inflammatory (palmitoleic^23^ and arachidonic acids^24^) and neurological (glutamate^25^ and cysteine-S-sulfate^26^) roles. Metabolites negatively associated with COVID-19, are involved in the urea cycle and nitric oxide (NO) synthesis pathway (proline, citrulline, and glutamine^27^). The NO synthesis pathway mediate responses to pro-inflammatory cytokines, macrophages, and neutrophils. Low levels of citrulline have been observed in patients with acute respiratory distress syndrome^28^ and can cause NO synthase uncoupling and decreased NO synthesis, which is important for vascular function and endothelial cell function^29^. It was recently suggested that therapeutic NO could be used to improve pulmonary vascular function in COVID-19^30^. Of the 17 metabolites associated with COVID-19 status, only glutamate was associated with disease severity. In addition, sex-specific correlations between immune responses and glutamate were observed by disease severity. Males that deteriorated from COVID-19 had positive correlations between glutamate and CD8 T cells, and a negative correlation with CD4 T cells. A previous study showed that higher innate immune cytokine levels are associated with clinical deterioration in females with COVID-19^2^, here we observed that IL6 was positively correlated with glutamate only in females that deteriorate. Increasing levels of glutamate have also been shown to decrease IL5 secretion^31^, and we observed a negative correlation between glutamate and IL5 in males that deteriorate and also in females that stabilize, furthermore, Eotaxin2 negatively correlates with glutamate in females that deteriorate. Similar to Eotaxin2, IL5 is also linked to eosinophil activation, therefore in females, glutamate may be important in regulating eosinophilia in COVID-19. Incidentally, KA is a glutamate receptor antagonist. Glutamate receptors are expressed on the surface of T cells and expression of these receptors is triggered by T cell activation^32^. Glutamate transporters have also been described in various immune cells^32^, therefore, this correlation could be therefore reflective of the actions of KA on glutamate levels and also immune cell responses to COVID-19^33^.

Because our study did not analyze non-COVID individuals exhibiting similar clinical symptoms to COVID-19 patients, it remains a possibility that elevated KA and KA:K may lead to elevated cytokines and more broadly mediate the inflammatory symptoms of other pathologies. This possibility, however, does not detract from our observations in COVID-19 patients or the potential of AhR as a therapeutic target in COVID-19.

In summary, we have identified serum metabolites associated with COVID-19 clinical course, immune response and sex-specific differences. Among these metabolites, perhaps the most salient discovery is the identification of KA as a metabolite associated with sex, age, increased disease severity, and elevated cytokine and chemokine levels. KA is a ligand for AhR, and when activated, AhR is a master regulator of immune responses and inflammation. Sex-specific agonism of AhR has yet to be reported in humans, but appears to be a prominent feature in COVID-19 disease, potentially underlying the cytokine storm and dampening of T cell activation. In addition, KA is known to dampen glutamate release^12^, and we observed lower levels of glutamate in patients that deteriorate compared to those that stabilize. Further investigation into the relevance of KA, KAT, and AhR activation in COVID-19 and the role of glutamate in clinical outcomes will be of utmost importance, particularly for understanding the sex-specific differences in immune response and patient outcomes. As we learn more about the impact of the metabolome on COVID-19 disease course, clinicians may find that modulating metabolite levels, either through enteral nutrition or targeted metabolic enzymes may alter disease trajectory.

## Data Availability

Untargeted metabolomics data, metabolomics protocols, and code is available on the MetaboLights data repository accession number MTBLS1987 (https://www.ebi.ac.uk/metabolights/). Clinical and immunological data is available from previous publication2.

## Methods

### Clinical biospecimens

Serum samples were collected from patients enrolled in the IMPACT study from Cohort A as described and stored at −20°C^2^. Cohort A consisted of 39 patients aged ≥ 18 years old that tested positive for SARS-CoV-2 by RT-PCR from nasopharyngeal and/or oropharyngeal swabs (females n = 22, males n = 17)^34^. Intersex individuals were not represented in this study. Prior to the serum collection, these patients were not in an intensive care unit, had not received tocilizumab and had not received high dose corticosteroids. Patients on hydroxychloroquine (n = 29) and remdesivir (n = 3) were not excluded. For control groups, we used 20 serum samples collected from COVID-19 uninfected health care workers working at the Yale-New Haven Hospital between April 2^nd^ and April 28^th^ 2020 who enrolled in the IMPACT study (females n = 10, males n = 10). The detailed demographics and clinical characteristics of these study participants and controls are shown in **Extended Data Table 1**.

### Immune markers and analysis of disease severity

An immune panel of markers for each patient was obtained and published in a previous study^2^. The patients were assessed with a locally developed clinical scoring system for disease severity^17^; 1: admitted and observed without supplemental oxygen, 2: required ≤ 3L supplemental oxygen via nasal canal to maintain SpO2 > 92%, 3: received tocilizumab, which per hospital treatment protocol required that the patient to require > 3L supplemental oxygen to maintain SpO2 > 92%, or, required > 2L supplemental oxygen to maintain SpO2 > 92% and had a high sensitivity C-reactive protein (CRP) > 70. 4: the patient required intensive care unit (ICU) level care, 5: the patient required intubation and mechanical ventilation. In relation to the WHO scoring, our clinical score 1, 2/3, 4, 5 largely correspond to WHO score 3, 4, 5, 6/7, respectively^35^. Detailed demographic information is available from^2^. For the patients who are 90-year-old or older, their ages were protected health information, and 90 was put as the surrogate value for the analyses. Individuals with active chemotherapy against cancers, pregnant patients, patients with background hematological abnormalities, patients with autoimmune diseases and patients with a history of organ transplantation and on immunosuppressive agents, were excluded from this study.

### Serum metabolite extraction

Serum samples (50 μL) were thawed and deactivated for COVID-19 in 150 μL acetone:methanol (50:50 v/v) for 60 min at room temperature. Control samples were treated in the exactly the same manner. To precipitate proteins, the samples were incubated for 2 hours at −20 °C, followed by centrifugation at 13,000 rpm (15,000× g) and 4 °C for 15 min. The resulting supernatant was removed and evaporated to dryness for 12 h using a vacuum concentrator (Thermo Fisher Scientific, Waltham, MA, USA). The dry extracts were then reconstituted in 100 μL of ACN:H_2_O (1:1, v/v), sonicated for 10 min, and centrifuged at 13,000 rpm (15,000× g) and 4 °C for 15 min, to remove insoluble debris. The supernatants were transferred to ultra performance liquid chromatography (UPLC) autosampler vials (Thermo Scientific, Waltham, MA, USA). A pooled quality control (QC) sample was prepared by mixing 5 μL of extracted solution from each sample into a similar UPLC vial. All the vials were then capped and stored at −80 °C prior to UPLC-mass spectrometry (MS) analysis.

### UPLC-MS-based metabolomics analysis

To comprehensively analyze the serum metabolome, both hydrophilic interaction chromatography (HILIC)-MS and reverse phase liquid chromatography (RPLC)-MS approaches were used. A UPLC system (H-Class ACQUITY, Waters Corporation, Milford, MA, USA), coupled to a quadrupole time-of flight (QTOF) (Xevo G2-XS QTOF, Waters Corporation, Milford, MA, USA), was used for MS data acquisition. A Waters ACQUITY UPLC BEH Amide column (particle size, 1.7μm; 100 mm (length) × 2.1 mm (i.d.)) and Waters ACQUITY UPLC BEH C18 column (particle size, 1.7 μm; 100 mm (length) × 2.1 mm (i.d.)) were used for the UPLC-based separation of metabolites. The column temperature was kept at 25 °C for HILIC-MS analysis and 30 °C for RPLC-MS analysis. The solvent flow rate was 0.5 mL/min, and the sample injection volume was 4 μL for HILIC-MS and RPLC in positive mode analysis, 2 μL for HILIC-MS in negative mode, and 6 μL for RPLC-MS negative mode. For HILIC-MS analysis, mobile phase A was 25 mM NH_4_OH and 25 mM NH_4_OAc in water, while the mobile phase B was acetonitrile, for both electrospray ionization (ESI), positive and negative mode, respectively. The linear gradient was set as follows: 0∼0.5 min: 95% B; 0.5∼7 min: 95% B to 65% B; 7∼8 min: 65% B to 40% B; 8∼9 min: 40% B; 9∼9.1 min: 40% B to 95% B; 9.1∼12 min: 95% B. For RPLC-MS analysis, the mobile phases A was 0.1% formic acid in H_2_O, while the mobile phases B was 0.1% formic acid in acetonitrile, respectively, for ESI+. Mobile phase A was 5 mM NH_4_OAc in H_2_O, while the mobile phases B was acetonitrile for ESI−. The linear gradient was set as follows: 0∼1 min: 1% B; 1∼8 min: 1% B to 100% B; 8∼10 min: 100% B; 10∼10.1 min: 100% B to 1% B; 10.1∼12 min: 1% B. Pooled samples were analyzed every eight injections during the UPLC-MS analysis to monitor the stability of the data acquisition and used for subsequent data normalization.

QTOF scan data (300 ms/scan; mass scan range 50–1000 Da) were initially acquired for each biological sample for metabolite quantification. Then, both DDA (data-dependent acquisition) data (QTOF scan time: 100 ms/scan, MSMS scan time 500 ms/scan, collision energy 20 eV, top 5 most intense ions were selected for fragmentation, exclude former target ions (4 s after 2 occurrences)) and MS^E^ data (low energy scan: 300 ms/scan, collision energy 6 eV; high energy scan: 300 ms/scan, collision energy 20 eV, mass scan range 25–1000 Da) were acquired for QC samples to enable metabolite identification. ESI source parameters on the Xevo GS-XS QTOF were set as the following: capillary voltage 1.8 kV, sampling cone 30 V, source temperature 100 °C, desolvation temperature 550 °C, cone gas flow 40 L/h, desolvation gas flow 900 L/h.

### UPLC-MS data processing

The raw MS data (.raw) were converted to mzML files using ProteoWizard MSConvert (version 3.0.6150, http://www.proteowizard.sourceforge.net/). The parameters of min SNR and min peak spacing were set as 0.1 for peak picking in ProteoWizard. The files were then processed in R (version 3.4.3), using the XCMS package for feature detection, retention time correction, and alignment^36^. The XCMS processing parameters were optimized and set as follows: mass accuracy for peak detection = 20 ppm; peak width c = (2, 30); snthresh = 6; bw = 10; mzwid = 0.015; minfrac = 0.5. The CAMERA package was used for subsequent peak annotation. The resulting data were normalized using the support vector regression algorithm in R, to remove an unwanted system error that occurred among intra- and inter-batches^37^. Initial metabolite identification was performed using the MetDNA algorithm^38^. Metabolites were further identified by matching retention time with an in-house metabolite standard library. In addition, metabolite identification was carried out by matching accurate mass and experimental MS/MS data against online databases (METLIN and HMDB).

### Multivariable logistic regression

Multivariable logistic regression analyses were performed on the R platform (version 3.4.3) using an R function “glm ()” The model for each metabolite was adjusted for age, BMI and sex to discover metabolites associated with COVID-19 disease. Levels of metabolites are log10 transformed ion intensity. *p* values were adjusted for multiple testing with Benjamini-Hochberg-based FDR using an R function “p.adjust ()”.

### Spearman correlation analysis

Spearman correlation analyses were performed on R platform (version 4.0.2) using an R package “psych”. Correlation coefficient R and *p* values were calculated using an R function “corr.test ()” Using previously defined interpretations of correlation coefficients, we used an |R| value of 0.5−1.0 to mark moderate-to-very high correlations^39^. Heatmaps were plotted using an R package “pheatmap”.

### Chord diagram

The chord diagrams were plotted on R platform (version 4.0.2) using an R package “circlize”. Correlations between metabolites and immune responses with R > 0.5 or < −0.5, and *p* value < 0.05 were plotted out.

### Gene expression analysis

Gene TPMs, subject phenotypes, and sample attributes data were downloaded from GTEX Portal (gtexportal.org, accession phs000424.v8.p2). After TPM values were transformed as log10(TPM+1), composite expression scores were calculated by adding individual expression values together. Patients who were 60 years or older were coded as “Older,” while patients 30 years or younger were coded as “Young.” After loading the expression data into R with the CePa package, Pearson correlation coefficients were calculated for pairs of target genes within each tissue of each sex, and data was visualized as a heatmap displaying the difference between male and female coefficients using the ComplexHeatmap package. Male-specific correlations were validated by scatter plots and linear regressions, which were generated using the ggplot2 R package.

## Data Availability and Code Availability

Untargeted metabolomics data, metabolomics protocols, and code is available on the MetaboLights data repository accession number MTBLS1987 (https://www.ebi.ac.uk/metabolights/). Clinical and immunological data is available from previous publication^2^. Data processing R code is available in Supplementary Information.

## Ethical statements

This study was approved by Yale Human Research Protection Program Institutional Review Boards (FWA00002571, Protocol ID. 2000027690). Informed consent was obtained from all enrolled patients and healthcare workers.

## Acknowledgements

We gratefully acknowledge the study participants for their time and commitment to the study. We thank all members of the clinical team at Yale-New Haven Hospital for their dedication and work which made this study possible. This work was in part supported by a gift from the Yale University Rapid Relief Fund, Women’s Health Research at Yale Pilot Project Program, Fast Grant from Emergent Ventures at the Mercatus Center, Mathers Foundation, the Beatrice Kleinberg Neuwirth Fund, and the Ludwig Family Foundation. IMPACT received support from the Yale COVID-19 Research Resource Fund. A.I. is an Investigator of the Howard Hughes Medical Institute. D.J.K. was a Paul and Daisy Soros Fellow and was supported by a grant from the National Cancer Institute (NCI) of the National Institutes of Health (NIH) (F30CA236466) and by a MSTP training grant from the NIH (T32GM007205, T32GM136651). S.A.K was supported by CTSA Grant Number UL1TR001863 from the National Center for Advancing Translational Science (NCATS), components of the NIH, and NIH roadmap for Medical Research, and the Lampman Research Fund in Yale Surgical Oncology. Support was also provided by the Beatrice Kleinberg Neuwirth Fund, Yale Schools of Public Health and Medicine, and NIH U19 AI08992 awarded to A.I.K.

## Author Contributions

All authors contributed to the data discussions and writing of this manuscript. Additional contributions: Y.C carried out the metabolomics mass spectrometry and data analysis, and prepared figures and tables, D.J.K carried out design of the gene expression analysis, performed the gene expression analysis and interpreted the results, T.T was involved in acquiring the biospecimens and experimental design, D.B validated statistical analysis design and reproduced the data independently, S.M. assisted with the biostatistics in the manuscript, N.J.W.R was involved in experimental design and figure design, A.C-M, A.J.M, M.C.M acquired the biospecimens and performed viral inactivation, B.I, J.K, C.L, T.M, J.O, J.S, P.W acquired immune cell data, A.I.K, contributed to data and clinical interpretation, S.A.K contributed to data interpretation and clinical discussions on data, A.I was involved in acquiring the biospecimens and experimental design, securing funds and supervising the study, C.H.J was involved in experiment design, data analysis, drafting the initial manuscript, securing funds and supervising the study. Y.C., D.B., C.H.J., and N.J.W.R are members of the COVID-19 Mass Spectrometry Coalition^40^.

## Yale IMPACT Research Team authors (in alphabetical order)

Tara Alpert, Kelly Anastasio, Michael H. Askenase, Maria Batsu, Santos Bermejo, Sean Bickerton, Anderson Brito, Kristina Brower, Molly L. Bucklin, Staci Cahill, Melissa Campbell, Yiyun Cao, Edward Courchaine, Rupak Datta, Giuseppe Deluliis, Charles Dela Cruz, Rebecca Earnest, Shelli Farhadian, Joseph Fauver, Renata Filler, John Fournier, Bertie Geng, Laura Glick, Nathan Grubaugh, Ryan Handoko, Christina Harden, Cole Jensen, Chaney Kalinich, William Khoury-Hanold, Lynda Knaggs, Maxine Kuang, Eriko Kudo, Sarah Lapidus, Joseph Lim, Melissa Linehan, Feimei Liu, Peiwen Lu, Alice Lu-Culligan, Maksym Minasyan, Amyn A. Malik, Anjelica Martin, Irene Matos, David McDonald, Maura Nakahata, Nida Naushad, Allison Nelson, Jessica Nouws, Angela Nuñez, Marcella Nunez-Smith, Abeer Obaid, Camila Odio, Ji Eun Oh, Saad B. Omer, Isabel M. Ott, Annsea Park, Hong-Jai Park, Xiaohua Peng, Mary Petrone, Sarah Prophet, Harold Rahming. Tyler Rice, Aaron Ring, Kadi-Ann Rose, Lorenzo Sewanan, Lokesh Sharma, Albert Shaw, Denise Shepard, Erin Silva, Mikhail Smolgovsky, Eric Song, Nicole Sonnert, Yvette Strong, Codruta Todeasa, Maria Tokuyama, Jordan Valdez, Sofia Velazquez, Arvind Venkataraman, Pavithra Vijayakumar, Chantal B.F. Vogels, Eric Y. Wang, Annie Watkins, Elizabeth B. White, Anne L. Wyllie, Yexin Yang.

## Additional Information

### Supplementary Information

This file contains Supplementary Table 1. Correlations between metabolites and immune markers in patients with COVID-19. Supplementary Table 2. Correlations between metabolites and immune markers in healthcare workers. The two tables include the calculated spearman correlation coefficient and *p* value. Supplementary Information contains data processing code for use in R.

### Extended Data Figures

**Extended Data Figure 1.**
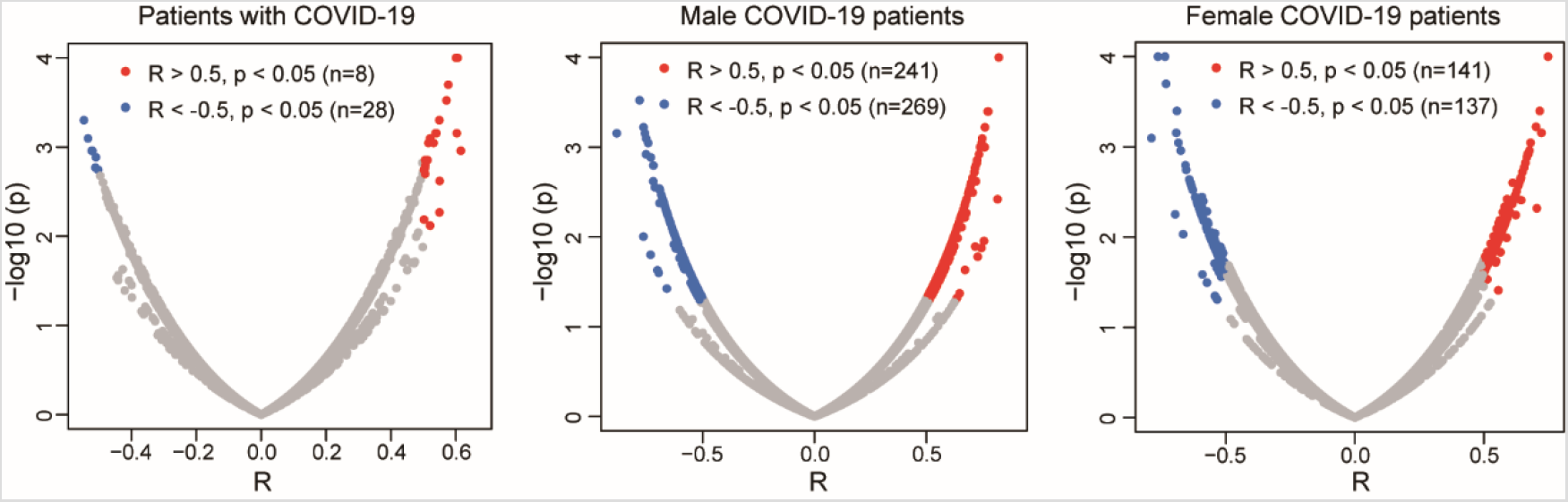
Scatter plots of correlation coefficient against −log10 (*p* value) between metabolites and immune markers in all patients with COVID-19, males with COVID-19, and females with COVID-19, respectively.

**Extended Data Figure 2.**
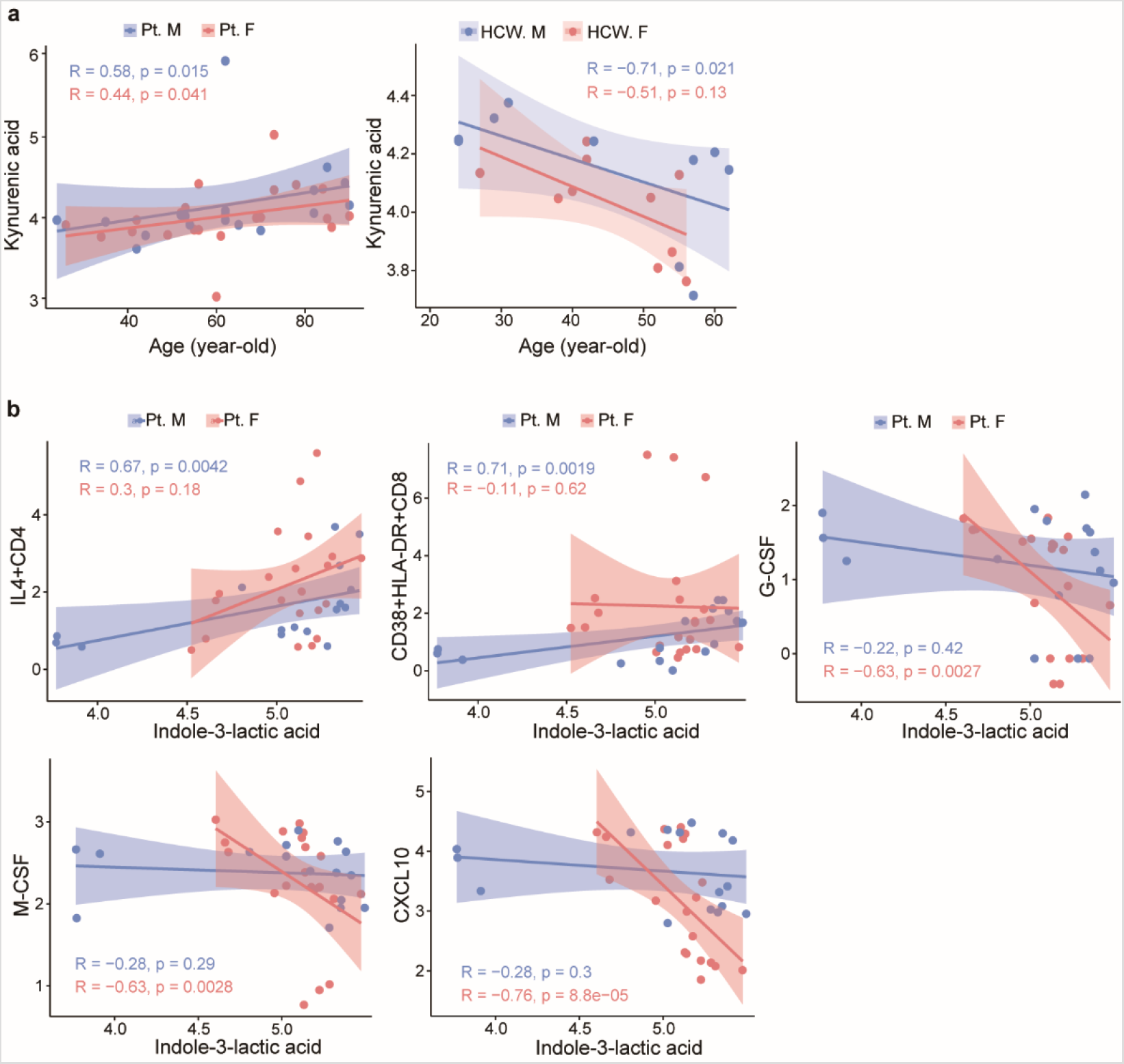
Correlations between metabolites and immune markers in patients with COVID-19 and healthcare workers stratified by sex. **a**, Correlation between age and kynurenic acid levels in patients with COVID-19 (left) and HCWs (right). **b**, Correlation between indole-3-lactic acid and IL4+CD4, CD38+HLA-DR+CD8, G-CSF, M-CSF and CXCL10 in males with COVID-19 and females with COVID-19, respectively. 95% confidence intervals (CIs) for the correlation coefficients were indicated as the shadowed area colored according to sex.

**Extended Data Figure 3.**
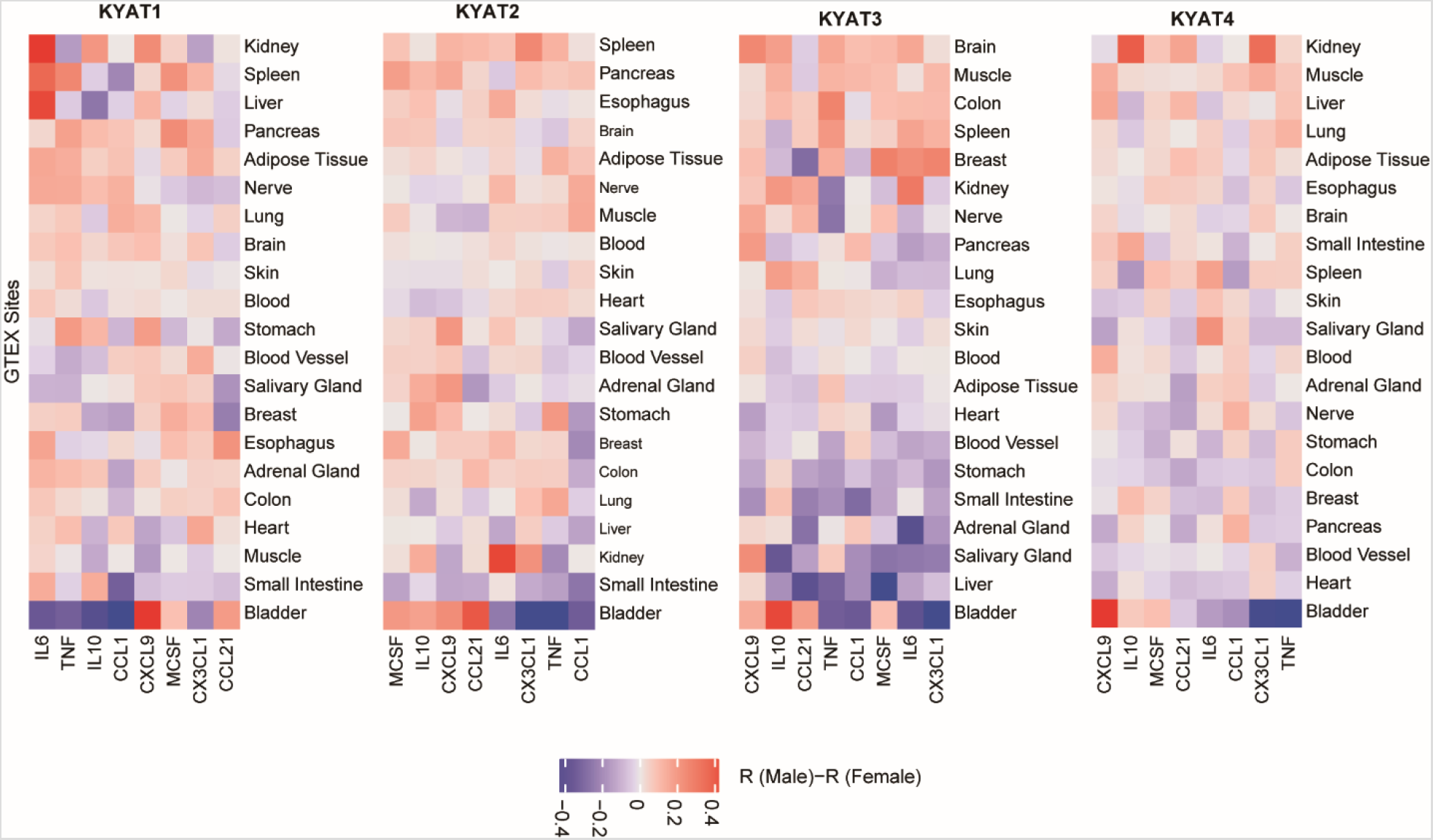
Correlations between KYAT gene expression and cytokines positively associated with either high KA or KA:K. Pearson correlation coefficients were calculated for gene pairs within the indicated tissue for each sex using GTEx data. Differences in the correlations (R_Male_–R_Female_) are presented as heatmaps, with red indicating a more positive correlation in males and blue indicating a more positive correlation in females (n = 729 males, 1914 females).

**Extended Data Figure 4.**
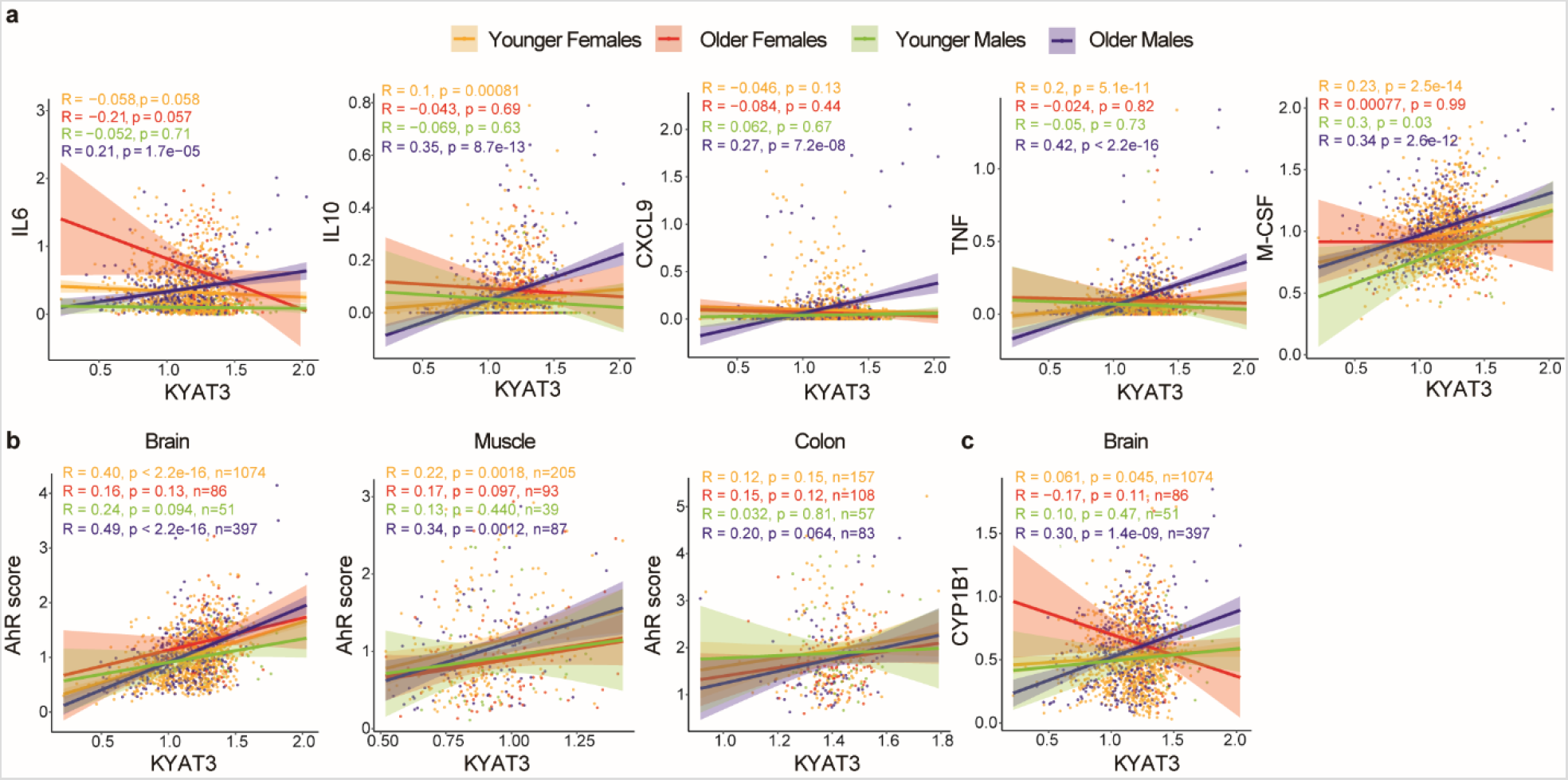
Correlations between KYAT3, immune markers, and AhR activation in younger and older individuals, stratified by sex. **a,** Correlations between *KYAT3* expression and *IL6, IL10, CXCL9, TNF*, and *M-CSF* in GTEx brain samples. **b**, Correlations between *KYAT3* and AhR activation score in brain, muscle and colon. **c**, Correlation between *KYAT3* and classic AhR target gene *CYP1B1* in brain.

### Extended Data table titles and footnotes

**Extended Data Table 1.** Demographic and clinical characteristics of patients with COVID-19 and healthcare workers

|  | Patients with COVID-19 (n=39) |  | Healthcare workers (n=20) |  |
| --- | --- | --- | --- | --- |
|  | Female n (%) | Male n (%) | Female n (%) | Male n (%) |
| **Total** | 22 (56) | 17 (44) | 10 (50) | 10 (50) |
| **Ethnicity\*** |  |  |  |  |
| Black/African American | 4 | 5 | 0 | 1 |
| White | 13 | 9 | 10 | 7 |
| Hispanic | 4 | 2 | 0 | 1 |
| **BMI\*** |  |  |  |  |
| < 18 | 1 | 0 | 0 | 0 |
| 18-24.9 | 6 | 4 | 3 | 3 |
| 25 – 29.9 | 5 | 7 | 4 | 2 |
| 30 – 34.9 | 8 | 3 | 0 | 3 |
| ≥35 | 2 | 3 | 3 | 2 |
| **Age – mean (SD)** | 60.2 (16.6) | 59.2 (17.8) | 45.7 (8.9) § | 44.2 (14.9) § |
| **Days from symptom onset – mean (SD)** | 12.3 (9.0) | 9.2 (5.5) | N/A | N/A |
| **Clinical Score – mean (SD)** | 1.27 (0.46) | 1.47 (0.62) | N/A | N/A |
| **On Hydroxychloroquine†** | 18 (81.8) | 11 (64.7) | N/A | N/A |
| **On Remdesivir†** | 2 (9.1) | 1 (5.9) | N/A | N/A |

N/A, not available for the data
\*Data included when available
†Status at first sample collection
§Student's *t*-test, $p < 0.05$ , comparing with patients with COVID-19

**Extended Data Table 2.**
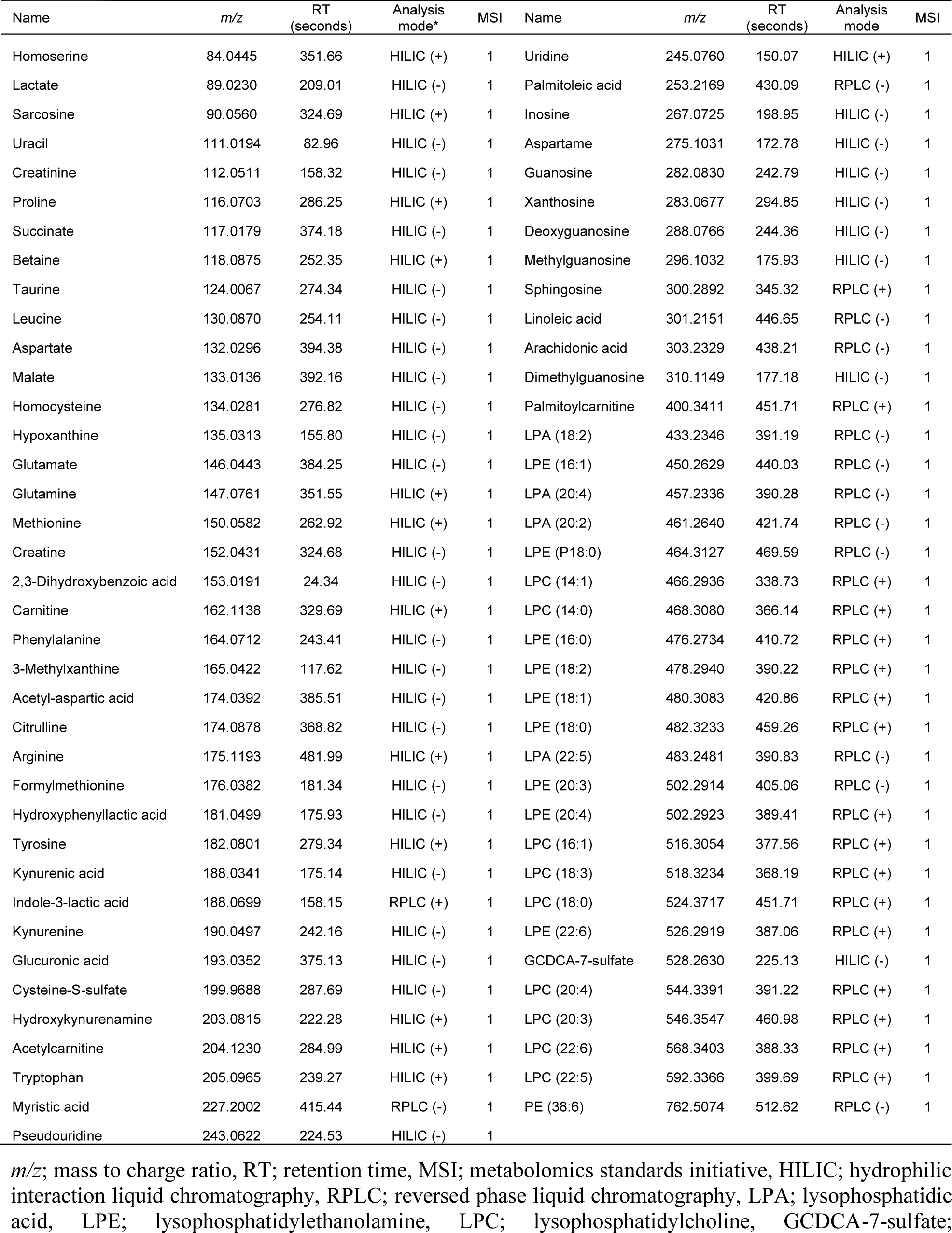
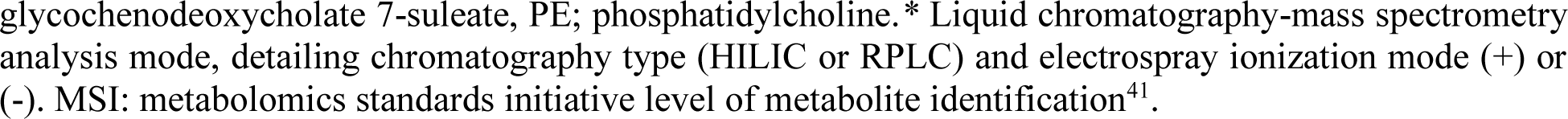
Positively identified metabolites from both COVID-19 patients and healthcare workers.

| Name | *m/z* | RT<br>(seconds) | Analysis<br>mode* | MSI | Name | *m/z* | RT<br>(seconds) | Analysis<br>mode | MSI |
| --- | --- | --- | --- | --- | --- | --- | --- | --- | --- |
| Homoserine | 84.0445 | 351.66 | HILIC (+) | 1 | Uridine | 245.0760 | 150.07 | HILIC (+) | 1 |
| Lactate | 89.0230 | 209.01 | HILIC (-) | 1 | Palmitoleic acid | 253.2169 | 430.09 | RPLC (-) | 1 |
| Sarcosine | 90.0560 | 324.69 | HILIC (+) | 1 | Inosine | 267.0725 | 198.95 | HILIC (-) | 1 |
| Uracil | 111.0194 | 82.96 | HILIC (-) | 1 | Aspartame | 275.1031 | 172.78 | HILIC (-) | 1 |
| Creatinine | 112.0511 | 158.32 | HILIC (-) | 1 | Guanosine | 282.0830 | 242.79 | HILIC (-) | 1 |
| Proline | 116.0703 | 286.25 | HILIC (+) | 1 | Xanthosine | 283.0677 | 294.85 | HILIC (-) | 1 |
| Succinate | 117.0179 | 374.18 | HILIC (-) | 1 | Deoxyguanosine | 288.0766 | 244.36 | HILIC (-) | 1 |
| Betaine | 118.0875 | 252.35 | HILIC (+) | 1 | Methylguanosine | 296.1032 | 175.93 | HILIC (-) | 1 |
| Taurine | 124.0067 | 274.34 | HILIC (-) | 1 | Sphingosine | 300.2892 | 345.32 | RPLC (+) | 1 |
| Leucine | 130.0870 | 254.11 | HILIC (-) | 1 | Linoleic acid | 301.2151 | 446.65 | RPLC (-) | 1 |
| Aspartate | 132.0296 | 394.38 | HILIC (-) | 1 | Arachidonic acid | 303.2329 | 438.21 | RPLC (-) | 1 |
| Malate | 133.0136 | 392.16 | HILIC (-) | 1 | Dimethylguanosine | 310.1149 | 177.18 | HILIC (-) | 1 |
| Homocysteine | 134.0281 | 276.82 | HILIC (-) | 1 | Palmitoylcarnitine | 400.3411 | 451.71 | RPLC (+) | 1 |
| Hypoxanthine | 135.0313 | 155.80 | HILIC (-) | 1 | LPA (18:2) | 433.2346 | 391.19 | RPLC (-) | 1 |
| Glutamate | 146.0443 | 384.25 | HILIC (-) | 1 | LPE (16:1) | 450.2629 | 440.03 | RPLC (-) | 1 |
| Glutamine | 147.0761 | 351.55 | HILIC (+) | 1 | LPA (20:4) | 457.2336 | 390.28 | RPLC (-) | 1 |
| Methionine | 150.0582 | 262.92 | HILIC (+) | 1 | LPA (20:2) | 461.2640 | 421.74 | RPLC (-) | 1 |
| Creatine | 152.0431 | 324.68 | HILIC (-) | 1 | LPE (P18:0) | 464.3127 | 469.59 | RPLC (-) | 1 |
| 2,3-Dihydroxybenzoic acid | 153.0191 | 24.34 | HILIC (-) | 1 | LPC (14:1) | 466.2936 | 338.73 | RPLC (+) | 1 |
| Carnitine | 162.1138 | 329.69 | HILIC (+) | 1 | LPC (14:0) | 468.3080 | 366.14 | RPLC (+) | 1 |
| Phenylalanine | 164.0712 | 243.41 | HILIC (-) | 1 | LPE (16:0) | 476.2734 | 410.72 | RPLC (+) | 1 |
| 3-Methylxanthine | 165.0422 | 117.62 | HILIC (-) | 1 | LPE (18:2) | 478.2940 | 390.22 | RPLC (+) | 1 |
| Acetyl-aspartic acid | 174.0392 | 385.51 | HILIC (-) | 1 | LPE (18:1) | 480.3083 | 420.86 | RPLC (+) | 1 |
| Citrulline | 174.0878 | 368.82 | HILIC (-) | 1 | LPE (18:0) | 482.3233 | 459.26 | RPLC (+) | 1 |
| Arginine | 175.1193 | 481.99 | HILIC (+) | 1 | LPA (22:5) | 483.2481 | 390.83 | RPLC (-) | 1 |
| Formylmethionine | 176.0382 | 181.34 | HILIC (-) | 1 | LPE (20:3) | 502.2914 | 405.06 | RPLC (-) | 1 |
| Hydroxyphenyllactic acid | 181.0499 | 175.93 | HILIC (-) | 1 | LPE (20:4) | 502.2923 | 389.41 | RPLC (+) | 1 |
| Tyrosine | 182.0801 | 279.34 | HILIC (+) | 1 | LPC (16:1) | 516.3054 | 377.56 | RPLC (+) | 1 |
| Kynurenic acid | 188.0341 | 175.14 | HILIC (-) | 1 | LPC (18:3) | 518.3234 | 368.19 | RPLC (+) | 1 |
| Indole-3-lactic acid | 188.0699 | 158.15 | RPLC (+) | 1 | LPC (18:0) | 524.3717 | 451.71 | RPLC (+) | 1 |
| Kynurenine | 190.0497 | 242.16 | HILIC (-) | 1 | LPE (22:6) | 526.2919 | 387.06 | RPLC (+) | 1 |
| Glucuronic acid | 193.0352 | 375.13 | HILIC (-) | 1 | GCDCA-7-sulfate | 528.2630 | 225.13 | HILIC (-) | 1 |
| Cysteine-S-sulfate | 199.9688 | 287.69 | HILIC (-) | 1 | LPC (20:4) | 544.3391 | 391.22 | RPLC (+) | 1 |
| Hydroxykynurenamine | 203.0815 | 222.28 | HILIC (+) | 1 | LPC (20:3) | 546.3547 | 460.98 | RPLC (+) | 1 |
| Acetylcarnitine | 204.1230 | 284.99 | HILIC (+) | 1 | LPC (22:6) | 568.3403 | 388.33 | RPLC (+) | 1 |
| Tryptophan | 205.0965 | 239.27 | HILIC (+) | 1 | LPC (22:5) | 592.3366 | 399.69 | RPLC (+) | 1 |
| Myristic acid | 227.2002 | 415.44 | RPLC (-) | 1 | PE (38:6) | 762.5074 | 512.62 | RPLC (-) | 1 |
| Pseudouridine | 243.0622 | 224.53 | HILIC (-) | 1 |  |  |  |  |  |

*m/z*; mass to charge ratio, RT; retention time, MSI; metabolomics standards initiative, HILIC; hydrophilic interaction liquid chromatography, RPLC; reversed phase liquid chromatography, LPA; lysophosphatidic acid, LPE; lysophosphatidylethanolamine, LPC; lysophosphatidylcholine, GCDCA-7-sulfate;
glycochenodeoxycholate 7-sulfate, PE; phosphatidylcholine.\* Liquid chromatography-mass spectrometry analysis mode, detailing chromatography type (HILIC or RPLC) and electrospray ionization mode (+) or (-). MSI: metabolomics standards initiative level of metabolite identification<sup>41</sup>.

**Extended Data Table 3.** Multivariable logistic regression analysis of metabolite levels from healthcare workers and COVID-19 patients.

| Metabolite | $\beta$ | *p* value | *q* value |
| --- | --- | --- | --- |
| Glutamate | 6.43 | 0.0018 | 0.0228 |
| Glutamine | -12.1 | 0.0014 | 0.0228 |
| Cysteine-S-sulfate | 3.67 | 0.0014 | 0.0228 |
| 3-Methylxanthine | -2.10 | 0.0015 | 0.0228 |
| Palmitoleic acid | 4.72 | 0.0008 | 0.0228 |
| Arachidonic acid | 6.35 | 0.0015 | 0.0228 |
| Tryptophan | -5.04 | 0.0044 | 0.0296 |
| Proline | -5.77 | 0.0049 | 0.0296 |
| Citrulline | -3.82 | 0.0051 | 0.0296 |
| Homoserine | -6.43 | 0.0041 | 0.0296 |
| 2,3-Dihydroxybenzoic acid | -3.11 | 0.0032 | 0.0296 |
| LPA (18:2) | -4.79 | 0.0050 | 0.0296 |
| LPE (22:6) | 4.74 | 0.0040 | 0.0296 |
| LPA (20:2) | -4.40 | 0.0058 | 0.0313 |
| Uracil | 5.04 | 0.0095 | 0.0467 |
| Myristic acid | 3.88 | 0.0100 | 0.0467 |
| LPC (14:0) | -7.23 | 0.0106 | 0.0467 |

Models were adjusted by age, sex, and BMI. *q*-values Benjamini-Hochberg adjusted *p* value. LPA; lysophosphatidic acid, LPE; lysophosphatidylethanolamine, LPC; lysophosphatidylcholine

